# Performance of Human and Computer-aided Evaluation of Digital Chest Radiography for Community-based Screening of Asymptomatic Tuberculosis

**DOI:** 10.64898/2026.04.29.26351560

**Authors:** Sarah Nyangu, Humphrey Mulenga, Simon C Mendelsohn, Tahlia Perumal, Michele Tameris, Tumelo Moloantoa, Stephanus T Malherbe, Firdows Noor, Justin Shenje, Nicolette Tredoux, Angelique Kany Kany Luabeya, Fernanda Maruri, Ravindre Panchia, Khuthadzo Hlongwane, Kim Stanley, Yuri F van der Heijden, Kate Hadley, Neil Martinson, Keertan Dheda, Al Leslie, Bernard Fourie, Gerhard Walzl, Thomas J Scriba, Timothy R Sterling, Mark Hatherill, the RePORT South Africa Study Team

**Affiliations:** South African Tuberculosis Vaccine Initiative, Institute of Infectious Disease & Molecular Medicine and Division of Immunology, Department of Pathology, University of Cape Town, Cape Town, South Africa; Vanderbilt Tuberculosis Center, Vanderbilt University Medical Center, Nashville, TN, USA; Perinatal HIV Research Unit (PHRU), University of the Witwatersrand, Johannesburg, South Africa; South African Medical Research Council Centre for Tuberculosis Research, Division of Immunology, Department of Biomedical Sciences, Faculty of Medicine and Health Sciences, Stellenbosch University, Cape Town, South Africa; The Aurum Institute, Johannesburg, South Africa; Centre for Lung Infection and Immunity, Division of Pulmonology, Department of Medicine and UCT Lung Institute & South African MRC/UCT Centre for the Study of Antimicrobial Resistance, University of Cape Town, Cape Town, South Africa; Africa Health Research Institute (AHRI), Durban, South Africa; Faculty of Infectious and Tropical Diseases, Department of Immunology and Infection, London School of Hygiene and Tropical Medicine, London, UK; Department of Medical Microbiology, University of Pretoria (UP) Faculty of Health Sciences, Pretoria, South Africa; South African MRC/UCT Centre for the Study of Antimicrobial Resistance, University of Cape Town, Cape Town, South Africa

**Keywords:** Asymptomatic, Tuberculosis, Household contacts, screening, digital CXR, CAD

## Abstract

**Background:** The World Health Organisation (WHO) recommends digital chest radiography (dCXR) with computer-aided detection (CAD) for tuberculosis (TB) screening of individuals >15 years of age.

**Methodology:** Adults (≥18 years) were enrolled (March 2021-December 2022) in South Africa into a community-based Screening Cohort (household contacts) and a facility-based Triage Cohort (symptomatic clinic attendees). Microbiologically-confirmed pulmonary TB required positive sputum culture and/or Xpert Ultra. Asymptomatic TB was diagnosed in participants without TB symptoms. dCXR were read by blinded human readers and qXR CAD (0.5 threshold; Qure.AI, India).

**Results:** dCXR from 1,353 participants (886 Screening Cohort; 467 Triage Cohort) were analysed. Microbiologically-confirmed TB occurred in 48 (5.4%) Screening Cohort [9 symptomatic (19%) and 39 asymptomatic (81%)]; and 116 (24.8%) Triage Cohort (all symptomatic) participants. dCXR sensitivity (human readers) for asymptomatic TB in the Screening Cohort was 56.4%, vs. 72.4% for symptomatic TB in the Triage Cohort (difference -16%; 95%CI -2.9 to -29.1); with specificities 94.1% and 81.2%, respectively. Corresponding qXR CAD sensitivities were 69.2% vs. 83.6% (difference -14.4%; 95%CI -26 to –2.8), with specificities 89.3% and 73.5%, respectively. The difference in dCXR sensitivity and specificity for asymptomatic TB between qXR CAD and human readers was 12.8% (95%CI -0.48 to 26.1) and -4.8% (95%CI -12.4 to 28.2), respectively.

**Conclusion:** Sensitivity of community-based dCXR screening for microbiologically-confirmed asymptomatic TB among household contacts was lower than for facility-based triage of symptomatic TB, but approached 70% with CAD. Neither human reader nor qXR CAD evaluation met WHO targets for a TB screening test (90% sensitivity; 80% specificity).

**Research in context:** *Evidence before this study:* The World Health Organisation (WHO) recommends digital chest radiography (dCXR) with computer-aided detection (CAD) for tuberculosis (TB) screening of individuals >15 years of age, based on data from prevalence surveys and facility-based studies. Performance data for community-based screening of asymptomatic TB are lacking. We searched PubMed for literature published in English between January 1, 2000, and November 1, 2025, for community-based, active case-finding studies of adolescents and adults aged 15 years and older that used dCXR CAD for asymptomatic TB screening. We used the following search terms: “Tuberculosis” AND (“asymptomatic” OR “subclinical”) AND (“computer aided diagnosis” OR “artificial intelligence”) AND “community-based screening” AND “chest radiography” AND (“diagnostic performance” OR “sensitivity”). We identified five studies reporting on microbiologically-confirmed asymptomatic TB and dCXR CAD performance. Three of five studies tested sputum only in those who were symptomatic and/or had abnormal CXR. One study did measure prevalence of asymptomatic TB by universal sputum testing of all participants, but did not report sensitivity and specificity for asymptomatic TB separately. One case-control study of CAD4TB (v7), which pooled data from five active case-finding cohorts, reported sensitivity of 61.4% and specificity of 86.7% for asymptomatic TB. However, the case-control design and inclusion of two cohorts using prevalence survey methodology and three cohorts enrolling high TB risk groups, two of which did not perform CXR on all participants, suggest potential for selection bias.

*Added value of this study:* We evaluated discriminatory performance of dCXR screening for asymptomatic TB among adult household contacts of TB patients, using human readers and qXR CAD (QURE.AI, India), in three communities in South Africa (Screening Cohort). Performance was benchmarked against that for symptomatic TB among adult clinic attendees (Triage Cohort), to enable comparison with traditional published approaches. All participants underwent universal sputum testing, regardless of symptom status or dCXR results. Sensitivity of human readers for asymptomatic TB in the Screening Cohort was 56.4%, compared to 72.4% for symptomatic TB in the Triage Cohort, with specificity 94.1% and 81.2%, respectively. The corresponding sensitivity of qXR CAD for asymptomatic TB, using the manufacturer’s 0.5 threshold score, was 69.2%, compared to 83.6% for symptomatic TB, with specificity 89.3% and 73.5%, respectively. The difference in dCXR sensitivity and specificity for asymptomatic TB between qXR CAD and human readers was 12.8% and -4.8%, respectively. The adjusted qXR threshold score (0.007) required to achieve 90% sensitivity for asymptomatic TB reduced specificity to 18.9%; and did not meet the WHO Target Product Profile (TPP) for a high sensitivity (90%), high specificity (80%) TB screening test.

*Implications of all the available evidence:* Sensitivity of community-based dCXR screening of household contacts for asymptomatic TB was low, compared to facility-based triage of symptomatic TB. Neither human reader nor qXR CAD evaluation of dCXR met the minimal WHO TPP for a high sensitivity (90%), high specificity (80%) TB screening test. Although dCXR CAD community screening would detect more than two-thirds of all people with previously undiagnosed, microbiologically-confirmed asymptomatic TB, the significant proportion of people with TB that would remain undetected, and untreated, might allow ongoing *Mycobacterium tuberculosis* transmission and hinder elimination efforts.

## Introduction

The World Health Organisation (WHO) has highlighted the importance of asymptomatic tuberculosis (TB) (1), occurring in individuals who do not have, recognise, or report symptoms of TB, but have microbiological evidence of *Mycobacterium tuberculosis* (Mtb) in the lungs or elsewhere. The natural history of asymptomatic TB is likely dynamic; modelling suggests that many individuals may regress without developing clinical disease, while a smaller proportion persist or progress over time (2). However, if infectious asymptomatic TB remains undetected for long periods, Mtb transmission might continue unchecked within communities and hinder elimination efforts (3, 4).

The burden of asymptomatic TB in national prevalence surveys is approximately 50% of all TB (5, 6). However, three community studies in South Africa found >80% of all TB to be asymptomatic using universal sputum testing, which highlights the ‘detection gap’ between universal testing and prevalence survey methodology, in which only positive symptom and/or chest radiograph (CXR) findings trigger diagnostic sputum sampling (7-9). Universal sputum testing includes asymptomatic individuals with negative CXR, who may have mild disease, providing a more accurate denominator to measure the impact of community screening.

We previously reported that CXR read by a single, non-expert reader detected only 56% of microbiologically-confirmed, asymptomatic TB among household contacts (HHC) in South Africa (7). Performance of digital CXR (dCXR) with computer-aided-detection (CAD) might be superior to non-expert human readers for detection of asymptomatic TB. dCXR with CAD was recommended by WHO in 2021 for TB screening of individuals >15 years of age, with expected sensitivity of 90% based on data from prevalence surveys and facility-based triage studies, which may over-estimate performance by selection for more severe disease (10, 11). Although some CAD software platforms appear marginally superior to others, the six platforms endorsed by WHO (CAD4TB v7, DrAid v2.4.4-6, Genki Edge v3.4.2, InferRead DR Chest v1.0.1.1, Insight CXR v3.1, and qXR v4.0) performed similarly to expert human readers in studies of symptomatic TB (11).

dCXR is readily available in many TB-endemic countries and CAD could be leveraged for mass screening in communities without access to expert radiographic readers. Given that countries are investing in dCXR screening programmes, it is important to understand how dCXR CAD performs for asymptomatic TB in community screening, in contrast to symptomatic TB in facility-based triage (7-9). The WHO Target Product Profile (TPP) for a high sensitivity, high specificity TB screening test requires minimum 90% sensitivity and 80% specificity (12). However, we lack sensitivity and specificity data for dCXR CAD detection of asymptomatic TB in an unselected population in which community prevalence is accurately defined by universal sputum testing. We evaluated the discriminatory performance of dCXR screening, using both human readers and CAD, among primarily asymptomatic household contacts of known TB patients in three South African communities. We benchmarked these findings against dCXR performance for symptomatic TB among clinic attendees, to enable comparison with traditional published approaches (13).

## Methods

### Study design

The Regional Prospective Observational Research for Tuberculosis (RePORT) South Africa network implemented a prospective observational study enrolling adults (≥18 years) between March 2021 to December 2022, into two parallel cohorts, using a single protocol, at six sites (7). The Screening Cohort (3 sites) included asymptomatic and symptomatic participants with recent TB exposure via household or other close contact (four or more hours per week), who were identified through index TB patient contact-tracing in the community. The Triage Cohort (3 sites) included symptomatic participants with possible TB presenting to fixed or mobile clinics.

### Procedures

At baseline, all participants underwent a symptom questionnaire, sputum collection, and dCXR evaluation. Participants reporting any of unexplained cough, fever, unintended weight loss, fatigue of lethargy, night sweats, pleuritic chest pain, and haemoptysis of any duration, were classified as symptomatic; those not reporting any symptom were classified as asymptomatic. A single spontaneous expectorated sputum was collected for acid-fast smear microscopy, Xpert Ultra (Cepheid, CA, USA) and liquid culture Mycobacteria Growth In Liquid (MGIT; BACTEC, Beckton Dickinson, NJ, USA). Participants unproductive of sputum were deemed sputum-negative; sputum induction was not performed. Microbiologically-confirmed prevalent pulmonary TB was defined by positive sputum Xpert-Ultra (excluding trace) and/or MGIT culture. The CXR was evaluated by a single reader (R1), usually the duty investigator at each site, with clinical training and experience in reviewing CXR features of TB, but not expert-level radiological expertise. R1 was not blind to participant-level clinical data, but was blind to the results of TB microbiological tests at the time of CXR review. A standardized Chest Radiographic Reading and Reporting System (CRRS) template for adults (14, 15), was adapted to report the radiological findings. The presence of one or more CXR features, including cavity, opacity, miliary pattern, tracheal shift, collapsed lung, or pleural effusion were deemed compatible with TB.

De-identified DICOM (Digital Imaging and Communications in Medicine) format dCXR were uploaded to a secure central server for evaluation by additional human readers (R2 and R3) with similar expertise to R1, who were blind to all participant-level clinical data, other dCXR reviews, and results of TB microbiological tests. R2 reviewed all available dCXR; R3 reviewed only dCXR with discordant R1 and R2 reviews. Final CXR classification (TB-positive or TB-negative) was determined by a two-thirds majority. Subsequently, de-identified dCXR images were uploaded to qXR (qXR software version 3, Qure.ai, India) for CAD evaluation. qXR provides a TB score on a probability scale of 0 to 1, with 1 being highly suggestive of TB. The qXR manufacturer’s recommended threshold for a CXR compatible with TB is 0.50.

### Statistical analysis

Statistical analysis was conducted using Stata-v18 and R-v2025.05.1. Descriptive statistics were computed as either mean and standard deviation (SD), or median and interquartile range (IQR), depending on distribution, or as frequencies and percentages for categorical variables. The Wilcoxon-Rank Sum and Kruskal-Wallis tests were used to compare continuous variables between two or more groups, respectively. Categorical variables were compared using the Fisher’s exact test. Statistical significance for all analyses was set at 0.05.

Area under the receiver operating characteristic curve (AUC), sensitivity and specificity of qXR scores were computed for discriminatory performance of microbiologically-confirmed asymptomatic TB and symptomatic TB, compared to no TB, in each cohort. 95% confidence intervals (CI) were computed using the DeLong method for AUC and exact binomial methods for diagnostic accuracy estimates (sensitivity/specificity).

The analysis comparing performance of qXR CAD and human readers was first computed using the manufacturer’s recommended 0.5 threshold score. Thereafter, alternative qXR thresholds were explored, benchmarked against the WHO high sensitivity TB screening test TPP (90% sensitivity, 80% specificity). Inter-rater reliability of human readers was tested using Cohen’s kappa statistical test (16).

### Ethical Considerations

This analysis was approved by the Human Research Ethics Committee of the University of Cape Town. The RePORT South Africa study protocol was approved by the institutional ethics committees of all participating sites. All participants provided written informed consent prior to participation.

## Results

A total of 1,353 of 1,732 enrolled participants (78.1%) had a dCXR available for reading by all three human readers and by qXR, including 467 from the Triage Cohort and 886 from the Screening Cohort (**Figure 1**). Screening and Triage Cohort participants differed in demographic characteristics, symptom status, likelihood of dCXR compatible with TB, and frequency of TB disease.

**Figure 1:**
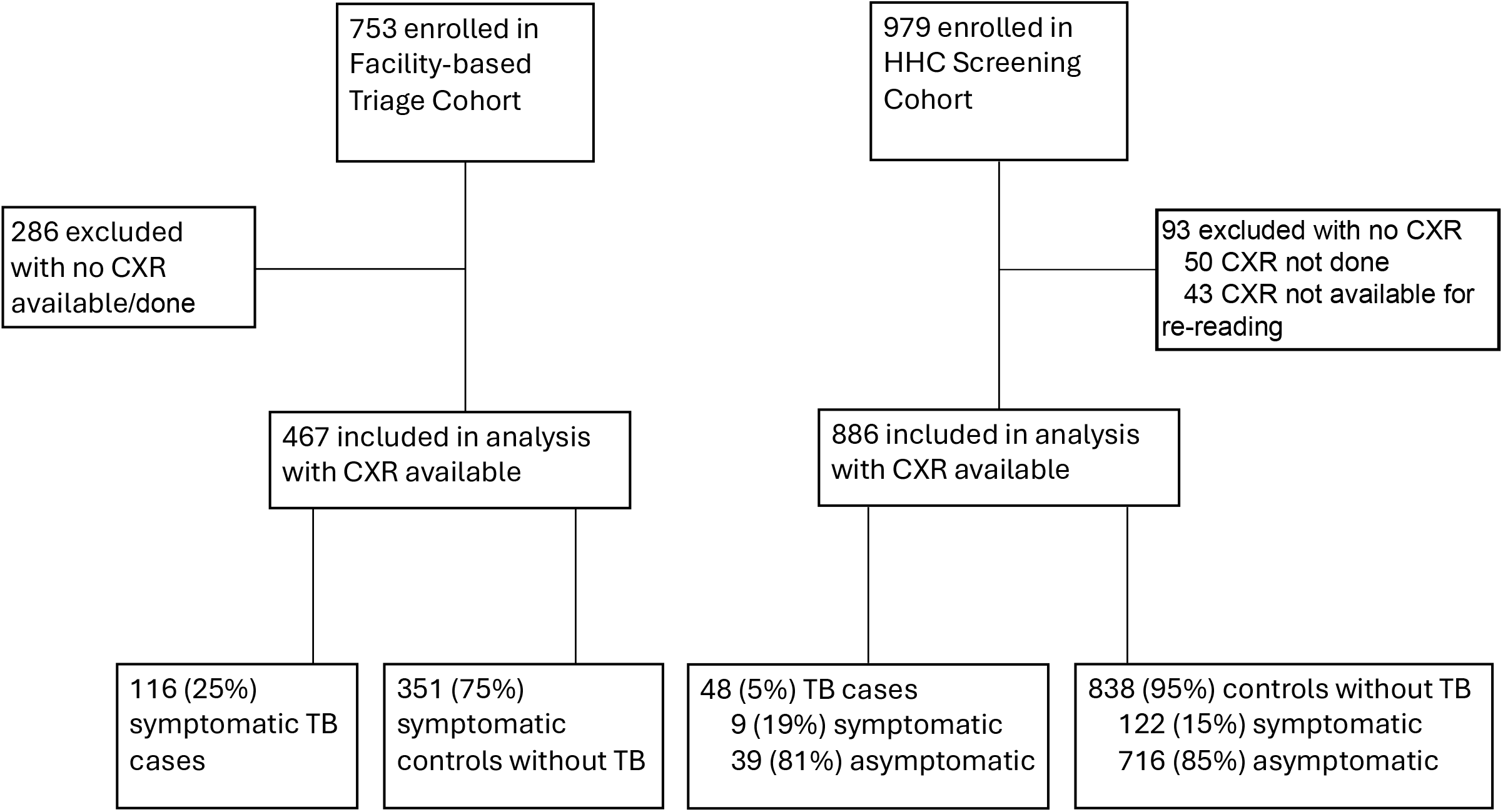
Flow of study participants.

In total, there were 168 participants with microbiologically-confirmed prevalent TB, including 48 of 886 (5.4%) from the Screening Cohort, of which 39 (81%) were asymptomatic and 9 (19%) symptomatic; and 116 of 467 (24.8%) from the Triage Cohort, all of whom were symptomatic. Among individuals without TB in the Screening Cohort, 716 of 838 (85%) participants were asymptomatic and 122 (15%) symptomatic. All 351 (75%) of 467 participants without TB in the Triage Cohort were symptomatic.

Compared to participants in the Screening Cohort, participants in the Triage Cohort were older (median 39.1 years versus 35.0 years, p<0.001); more likely to be male (53.9% versus 36.2%, p<0.001); living with HIV (27.6% versus 16.8%, p<0.001); or previous smokers (10.2% versus 5.3%, p<0.001); and to have had prior TB (35.3% versus 20.4%, p<0.001). Symptomatic participants in the Triage Cohort had a higher proportion of dCXR compatible with TB, compared to participants in the Screening Cohort, whether reported by human readers (32.1% versus 10.1%, p<0.001) or qXR (40.7% versus 14.8%, p<0.001; **Table 1**).

**Table 1:** Participant demographic and clinical characteristics stratified by cohort and symptom status.

| Characteristic | Triage Cohort | Screening Cohort |  |  | P value* |
| --- | --- | --- | --- | --- | --- |
|  | Symptomatic<br>N=467 | Asymptomatic<br>N=755 | Symptomatic<br>N=131 | All<br>N=886 |  |
| **Age, median (IQR)** | 39.1 (30.6–48.0) | 33.7 (24.9–46.6) | 43.1 (32.4–53.7) | 35.0 (25.8–48.2) | <0.001 |
| **Male sex, n (%)** | 252 (54.0) | 269 (35.6) | 52 (39.7) | 321 (36.2) | <0.001 |
| **Ethnicity, n (%)** |  |  |  |  | <0.001 |
| Black African | 309 (66.2) | 306 (40.5) | 96 (73.3) | 402 (45.4) |  |
| Cape mixed ancestry | 151 (32.3) | 446 (59.1) | 35 (26.7) | 481 (54.3) |  |
| Other | 7 (1.5) | 3 (0.4) | 0 | 3 (0.34) |  |
| **CXR compatible with TB, n (%)** |  |  |  |  |  |
| Reader 1 | 213 (45.6) | 99 (13.1) | 28 (21.4) | 127 (14.3) |  |
| Reader 2 | 145 (31.0) | 78 (10.3) | 28 (21.4) | 106 (12.0) |  |
| Reader 3 | 19 (4.1) | 16 (2.1) | 4 (3.1) | 20 (2.3) |  |
| **Reader consensus (2/3 reads)** | 150 (32.1) | 64 (8.5) | 25 (19.1) | 89 (10.0) | <0.001 |
| **qXR** | 190 (40.7) | 102 (13.5) | 29 (22.1) | 131 (14.8) | <0.001 |
| **Median qXR TB score (IQR)** |  |  |  |  |  |
| TB cases | 0.95 (0.82–0.97) | 0.88 (0.15–0.97) | 0.97 (0.91–0.98) | 0.91 (0.21–0.97) | 0.060 |
| No TB | 0.05 (0.02–0.52) | 0.02 (0.01–0.05) | 0.05 (0.01–0.27) | 0.02 (0.01–0.06) | <0.001 |
| **HIV positive, n (%)** | 129 (27.6) | 108 (14.3) | 41 (31.3) | 149 (16.8) | <0.001 |
| **Symptom positive, n (%)** | 467 (100) | 0 | 131 (100) | 131 (14.8) | <0.001 |
| Cough | 429 (91.9) | 0 | 96 (73.3) | 96 (10.8) |  |
| Night sweats | 296 (63.4) | 0 | 34 (26.0) | 34 (3.8) |  |
| Weight loss | 242 (51.8) | 0 | 54 (41.2) | 54 (6.1) |  |
| Pleuritic chest pain | 208 (44.5) | 0 | 33 (25.2) | 33 (3.7) |  |
| Fatigue | 185 (39.6) | 0 | 23 (17.6) | 23 (2.6) |  |
| Fever | 115 (24.6) | 0 | 31 (23.7) | 31 (3.5) |  |
| Haemoptysis | 35 (7.5) | 0 | 4 (3.1) | 4 (0.5) |  |
| **Smoking history, n (%)** |  |  |  |  | 0.001 |
| Current smoker | 256 (54.8) | 407 (53.9) | 67 (51.2) | 474 (53.5) |  |
| Former smoker | 48 (10.3) | 36 (4.8) | 11 (8.4) | 47 (5.3) |  |
| Never smoked | 162 (34.7) | 312 (41.3) | 53 (40.5) | 365 (41.2) |  |
| Missing<sup>‡</sup> | 1 (0.2) |  |  |  |  |
| **Previous TB disease, n (%)** | 165 (35.3) | 143 (18.9) | 38 (29.0) | 181 (20.4) | <0.001 |
| **Current TB disease, n (%)** | 116 (24.8) | 39 (5.2%) | 9 (6.9%) | 48 (5.4) | <0.001 |
| **Sputum microbiological results, n (%)<sup>#</sup>** |  |  |  |  |  |
| Positive smear microscopy | 81 (17.3) | 21 (2.8) | 6 (4.6) | 27 (3.0) | <0.001 |
| Positive Xpert MTB/RIF Ultra | 116 (24.8) | 36 (4.8) | 9 (6.9) | 45 (5.1) | <0.001 |
| Positive MGIT culture | 102 (21.8) | 32 (4.2) | 8 (6.1) | 40 (4.5) | 0.002 |
| Microbiological reference standard | 116 (24.8) | 39 (5.2) | 9 (6.9) | 48 (5.4) | <0.001 |
| **Median MGIT culture time to positivity, days (IQR)** | 8.5 (6–14) | 11 (6–20) | 12.5 (8.5–18.3) | 11 (6.3–18.8) | 0.042 |
| **Xpert MTB/RIF Ultra semi-quantitative result, n (%)<sup>‡</sup>** |  |  |  |  | 0.14 |
| High | 34 (7.3) | 8 (1.1) | 2 (1.5) | 10 (1.1) |  |
| Medium | 19 (4.1) | 4 (0.5) | 2 (1.5) | 6 (0.7) |  |
| Low | 33 (7.1) | 8 (1.1) | 2 (1.5) | 10 (1.1) |  |
| Very low | 14 (2.9) | 10 (1.3) | 2 (1.5) | 12 (1.4) |  |
| Trace | 15 (3.2) | 6 (0.8) | 1 (0.8) | 7 (0.8) |  |
| Missing<sup>‡</sup> | 1 (0.2) |  |  |  |  |

<sup>‡</sup>Not categorised: missing one person in the smoking group, Xpert semi-quantitative.
\*P values are Fisher's exact test for comparison of Triage Cohort (n=467) and Screening Cohort (n=886).

Sensitivity of dCXR (human readers) for asymptomatic TB in the Screening Cohort was low (56.4%), with 94.1% specificity; whereas sensitivity was high (88.9%) for symptomatic TB in the Screening Cohort, with 86.1% specificity. In the Triage Cohort, sensitivity was moderate for symptomatic TB (72.4%), with 81.2% specificity (**Table 2**). dCXR sensitivity (human readers) for asymptomatic TB in the Screening Cohort was significantly less than for symptomatic TB in the Triage Cohort (difference -16%; 95%CI -2.9 to -29.1).

**Table 2:** Diagnostic performance of qXR vs human readers in the Triage and Screening Cohorts.

|  | Triage Cohort<br>% (95% CI) |  | Screening Cohort<br>% (95% CI) |  |  |  |  |  |
| --- | --- | --- | --- | --- | --- | --- | --- | --- |
|  | Symptomatic |  | All |  | Symptomatic |  | Asymptomatic |  |
|  | Sensitivity | Specificity | Sensitivity | Specificity | Sensitivity | Specificity | Sensitivity | Specificity |
| **Human Reader 1** | **82.8**<br>(74.6–89.1) | **66.7**<br>(61.5–71.6) | **62.5**<br>(47.4–76.0) | **88.2**<br>(85.8–90.3) | **88.9**<br>(51.8–99.7) | **83.6**<br>(75.8–89.7) | **56.4**<br>(39.6–72.2) | **89.0**<br>(86.5–91.2) |
| **Human Reader 2** | **71.6**<br>(62.4–79.5) | **82.3**<br>(77.9–86.2) | **62.5**<br>(47.4–76.0) | **90.9**<br>(88.8–92.8) | **77.8**<br>(40.0–97.2) | **82.8**<br>(74.9–89.0) | **59.0**<br>(42.1–74.4) | **92.1**<br>(89.8–93.9) |
| **Human Reader Consensus (2/3)** | **72.4**<br>(63.3–80.3) | **81.2**<br>(76.7–85.1) | **62.5**<br>(47.4–76.0) | **93.0**<br>(91.0–94.6) | **88.9**<br>(51.8–99.7) | **86.1**<br>(78.6–91.7) | **56.4**<br>(39.6–72.2) | **94.1**<br>(92.2–95.7) |
| **qXR 0.5 threshold** | **83.6**<br>(75.6–89.8) | **73.5**<br>(68.6–78.0) | **72.9**<br>(58.2–84.7) | **88.3**<br>(86.0–90.4) | **88.9**<br>(51.8–99.7) | **82.8**<br>(74.9–89.0) | **69.2**<br>(52.4–83.0) | **89.3**<br>(86.8–91.4) |
| **qXR threshold adjusted to match human reader consensus (2/3) sensitivity** | **72.4**<br>(63.3–80.3)<br>[threshold 0.86] | **90.3**<br>(0.91–0.94)<br>[threshold 0.86] | **62.5**<br>(47.4–76.0)<br>[threshold 0.775] | **92.9**<br>(0.88–0.92)<br>[threshold 0.775] | **88.9**<br>(51.8–99.7)<br>[threshold 0.885] | **89.3**<br>(0.93–0.95)<br>[threshold 0.885] | **56.4**<br>(39.6–72.2)<br>[threshold 0.775] | **94.1**<br>(0.88–0.92)<br>[threshold 0.775] |
| **qXR threshold adjusted to match WHO TPP minimal sensitivity (90%)\*** | **90%**<br>[threshold 0.13] | **61.9%**<br>(56.5–67.3) | **90%**<br>[threshold 0.018] | **50.2 %**<br>(46.8–53.6) | **#89%**<br>[threshold 0.885] | **89.3 %**<br>(82.5–94.2) | **90%**<br>[threshold 0.007] | **18.9%**<br>(16.1–21.9) |

\*Adjusted qXR CAD threshold required to match WHO TPP sensitivity (90%)

qXR CAD scores differed by cohort, symptom status and TB status. In the Screening Cohort, median qXR CAD scores were higher among individuals with asymptomatic TB (0.88), compared to those without TB (0.02; p<0.001), but marginally lower than among individuals with symptomatic TB (0.97; p=0.070). Median qXR scores were marginally higher among those with symptomatic TB in the Triage Cohort (0.95) compared to those with asymptomatic TB in the Screening Cohort (0.88; p=0.060; **Figure 2**). qXR scores among participants without TB were also higher in the Triage Cohort (median 0.05) vs. the Screening Cohort (median 0.02; p=0.001).

**Figure 2:**
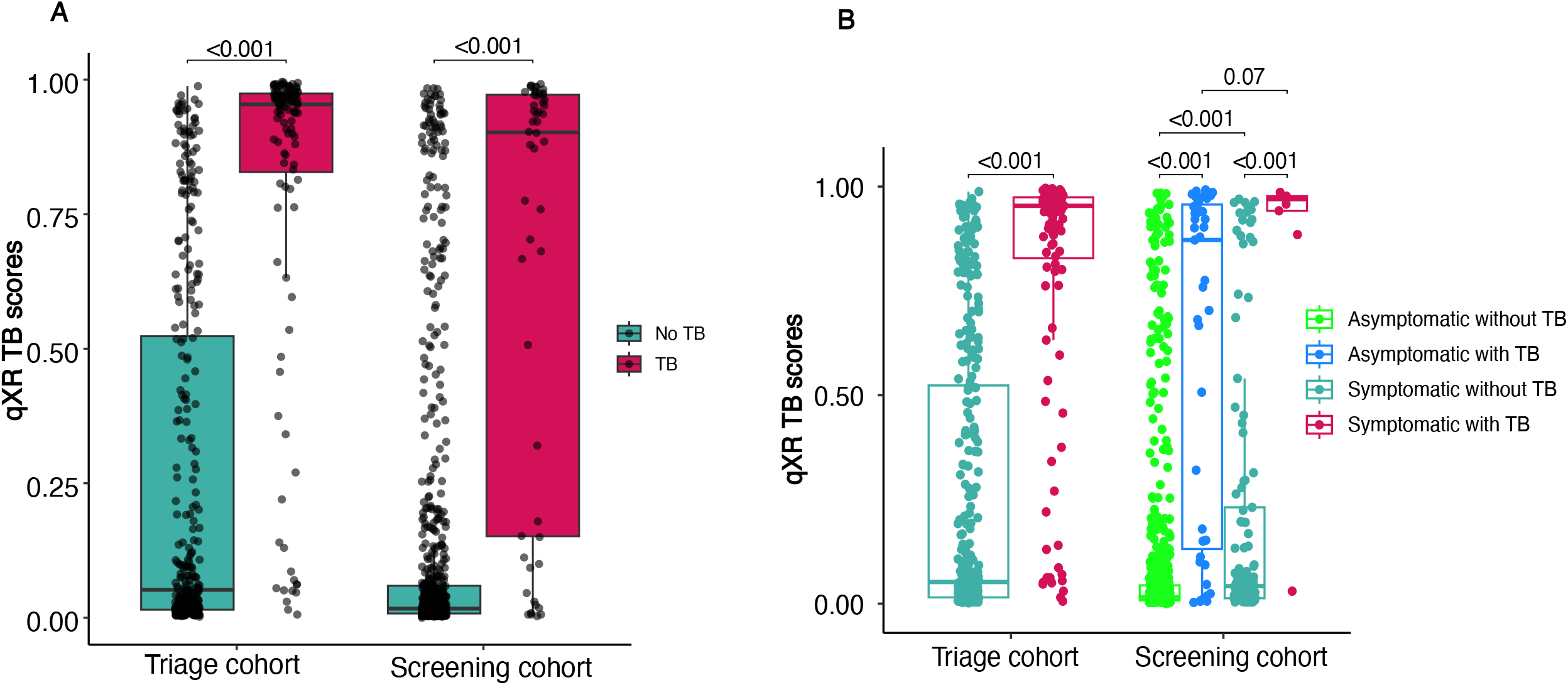
qXR TB CAD scores for the Triage Cohort and Screening Cohort. (**A**) Box-and-whiskers plots comparing qXR TB scores in the Triage and Screening Cohorts among TB cases and controls without TB, and (**B**) stratified by symptom status. Each dot represents one participant. Boxes depict the IQR, the midline represents the median, and the whiskers indicate the IQR ± (1.5 × IQR). P values of between-group differences were computed by Wilcoxon Rank-Sum Test.

Overall performance of qXR CAD in the Screening Cohort for discrimination of all participants with and without TB (AUC 0.84, 95%CI 0.77–0.91; p=0.21; **Figure 3A)**, was similar for discrimination of asymptomatic participants with and without TB (AUC 0.83, 95%CI 0.75–0.91; p=0.18; **Figure 3B)**, and in the Triage Cohort for discrimination of symptomatic participants with and without TB (AUC 0.89, 95%CI 0.86–0.93). Discriminatory performance of dCXR with qXR CAD was superior for persons without prior TB (AUC 0.93, 95%CI 0.89–0.96), compared to those with prior TB (AUC 0.82, 95%CI 0.72–0.90) in the Triage Cohort (p=0.010; **Figure 4**). No difference in performance was observed by prior TB status in the Screening Cohort. Performance of qXR was similar for persons with and without HIV in both cohorts (**Supplementary Figure 2**).

**Figure 3:**
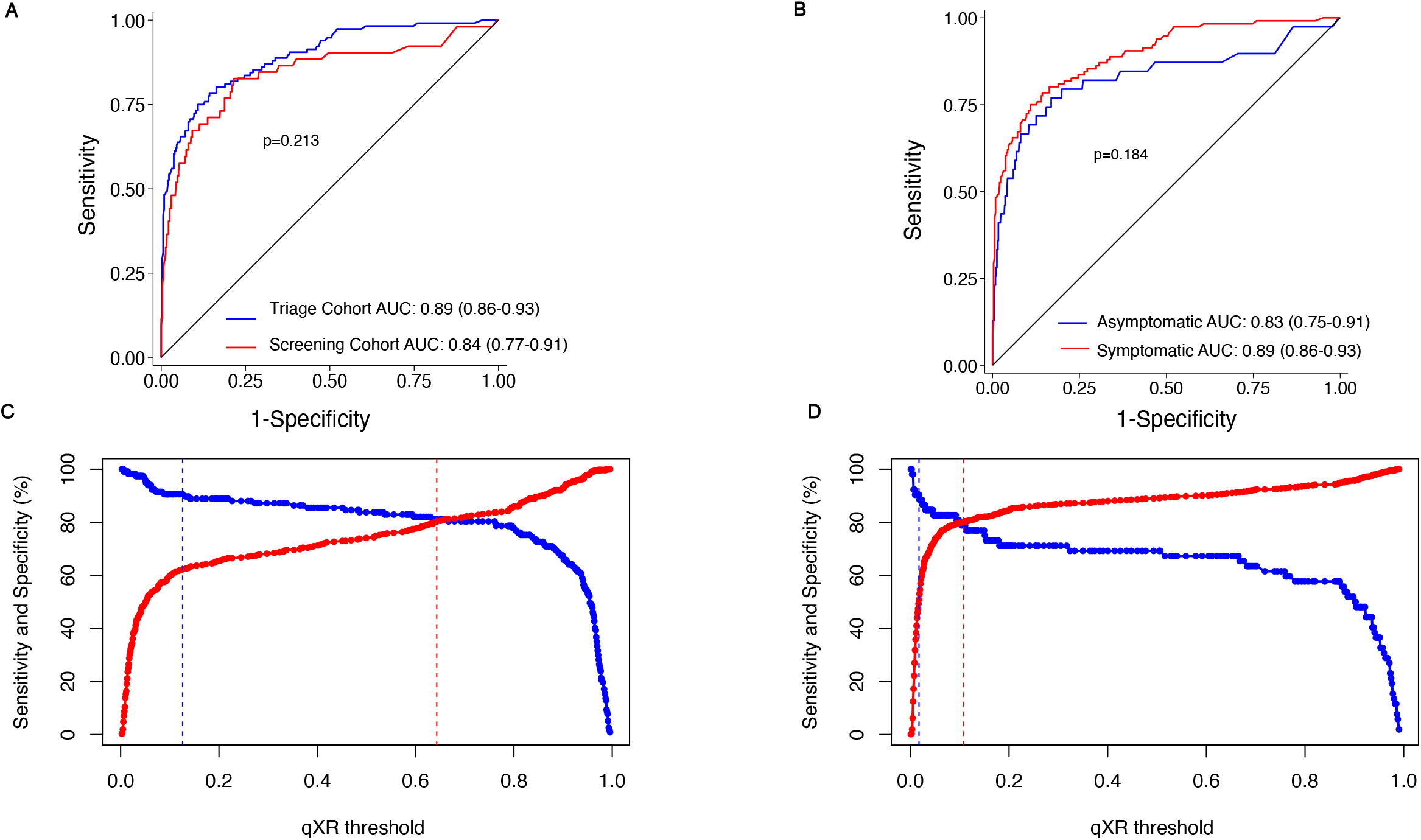
Performance of qXR for diagnosing TB in the Triage and Screening cohorts. (**A**)Receiver operating characteristic (ROC) curve with area under the curve (AUC) and 95% confidence intervals (CI) for qXR differentiating TB cases from controls without TB in the Triage (n=116 TB cases and n=351 controls without TB) and Screening (n=48 TB cases and n=838 controls without TB) Cohorts. The p-value shown is derived from the DeLong test comparing AUCs between the two cohorts. (**B**)ROC curve with AUC and 95% CI for qXR differentiating symptomatic TB cases (n=116) from symptomatic controls without TB (n=351) in the Triage Cohort; and asymptomatic TB (n=48) from all controls without TB (n=838) in the Screening Cohort. AUC with 95% CI are shown separately for symptom-defined sub-groups. The p-value shown is derived from the DeLong test comparing AUCs between symptomatic and asymptomatic groups. (**C**)and (**D**) show qXR sensitivity (solid blue line) and specificity (solid red line) plotted against the qXR score threshold for diagnosing TB in (**C**) symptomatic participants in the Triage Cohort; and (**D**) asymptomatic participants in the Screening Cohort. The dashed vertical red line represents the Youden’s Index qXR threshold. The dashed vertical blue line represents the qXR threshold at which a sensitivity of 90% (per the WHO TPP benchmark for a screening test) is attained, with corresponding specificity indicated.

**Figure 4:**
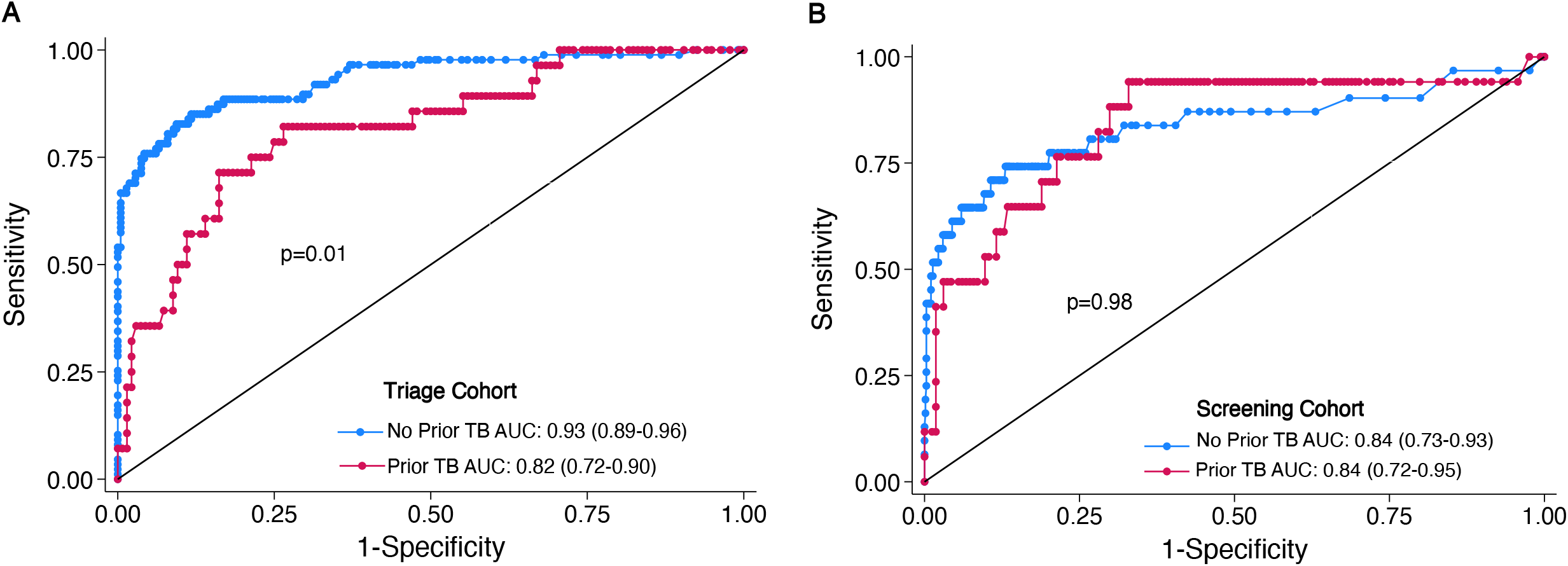
Performance of qXR for diagnosing TB in the Triage and Screening Cohorts, stratified by history of prior TB. Receiver operating characteristic (ROC) curves for qXR differentiating TB cases from controls without TB in the (**A**) Triage and (**B**) Screening Cohorts, stratified by prior TB history. Area under the curve (AUC) with 95% confidence intervals is shown separately for sub-groups with and without a history of prior TB (Triage Cohort: n=165 with, and n=302 without prior TB history; Screening Cohort: n=181 with, and n=705 without prior TB history). The p-values shown are derived from the DeLong test comparing AUCs between sub-groups with and without a history of prior TB in each cohort.

Sensitivity of qXR CAD at the manufacturer’s 0.5 threshold score was moderate for all TB in the Screening Cohort (72.9%), with 88.3% specificity; and high for symptomatic TB in the Triage Cohort (83.6%), with 73.5% specificity. When disaggregated by symptom status in the Screening Cohort, sensitivity for asymptomatic TB was 69.2%, with 89.3% specificity, whereas sensitivity for symptomatic TB was 88.9%, with 82.8% specificity (**Table 2**). qXR CAD sensitivity for asymptomatic TB in the Screening Cohort was significantly less than sensitivity for symptomatic TB in the Triage Cohort (difference -14.4%; 95%CI -25.9 to –2.8).

The difference in dCXR sensitivity and specificity for asymptomatic TB between qXR CAD and human readers was 12.8% (95%CI -0.48 to 26.1) and -4.8% (95%CI -12.4 to 28.2), respectively.

The qXR threshold score at Youden’s index (approximately 80% sensitivity and 80% specificity) was markedly lower for asymptomatic TB in the Screening Cohort (0.13), compared to symptomatic TB in the Triage Cohort (0.65; **Figure 3C-D**). When performance of dCXR for asymptomatic TB in the Screening Cohort was compared to the WHO TPP (12), neither human readers (56.4% sensitivity; 94.1% specificity), nor qXR CAD at the manufacturer’s threshold score (69.2% sensitivity; 89.3% specificity), approached the minimum criteria for a high sensitivity (90%) and high specificity (80%) screening test. However, although neither human readers nor qXR CAD met these minimum criteria for symptomatic TB in the Triage Cohort, both human readers and qXR CAD approached the minimum TPP criteria for symptomatic TB in the Screening Cohort.

Lowering the qXR CAD threshold score from the manufacturer’s 0.5 threshold to 0.018, in order to achieve the WHO TPP 90% sensitivity benchmark in the Screening Cohort, also reduced specificity to 50.2% (**Table 2**). The qXR threshold required to achieve 90% sensitivity for asymptomatic TB in the Screening Cohort was even lower (0.007), resulting in only 18.9% specificity.

Inter-rater agreement between human readers and qXR was substantial for asymptomatic TB in both the Triage and Screening Cohorts; and was almost perfect for symptomatic TB within the Screening Cohort (**Table 3A-B**).

**Table 3A:** Agreement between human readers and qXR for Triage and Screening Cohorts.

|  | Triage Cohort |  |  |  | Screening Cohort |  |  |  |
| --- | --- | --- | --- | --- | --- | --- | --- | --- |
|  | Agreement | Expected agreement | Kappa | P value | Agreement | Expected agreement | Kappa | P value |
| TB cases | 88.7% | 65.1% | 0.679 | <0.000 | 85.4% | 55.7% | 0.671 | <0.001 |
| No TB | 84.3% | 64.7% | 0.557 | <0.000 | 93.7% | 83.1% | 0.625 | <0.001 |
| **Overall** | **85.4%** | **53.3%** | **0.688** | **<0.000** | **93.2%** | **78.1%** | **0.690** | **<0.001** |

**Table 3B.**
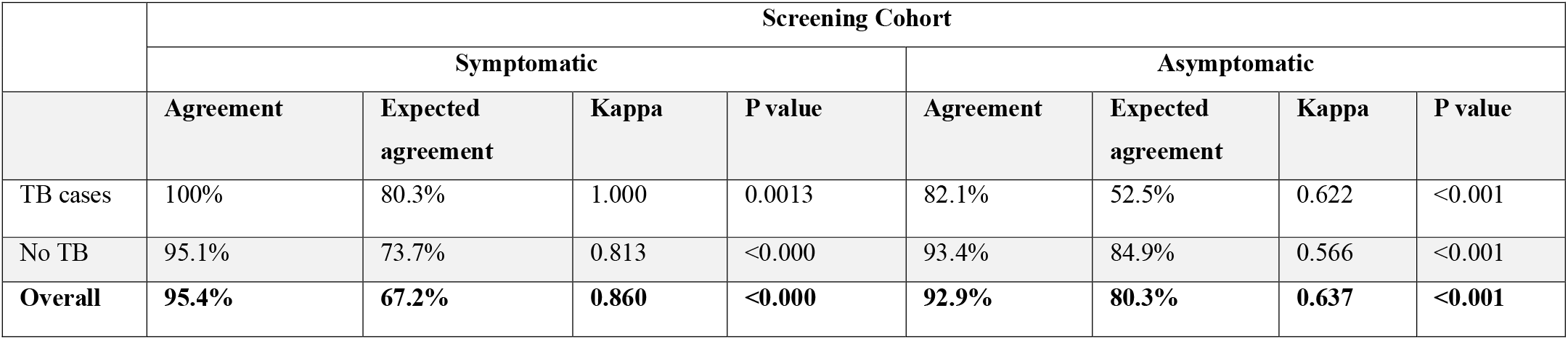
Agreement between human readers and qXR for symptomatic and asymptomatic persons in the Screening Cohort.

## Discussion

We demonstrated that sensitivity of community-based dCXR screening for microbiologically-confirmed, asymptomatic pulmonary TB among household contacts was lower than for symptomatic TB amongst clinic attendees, when dCXR were evaluated by multiple, non-expert human readers. dCXR evaluation by qXR showed a wide distribution of CAD scores for individuals with asymptomatic TB, which considerably overlapped those without TB. Although sensitivity of qXR CAD for asymptomatic TB (69.2%) appeared marginally higher than human readers (56.4%), with a corresponding minor decrease in specificity, neither dCXR evaluation approach met the WHO TPP for a high-sensitivity (90%), high-specificity (80%) TB screening test (12). Attempts to increase qXR CAD sensitivity by artificially lowering the threshold score resulted in unacceptably low specificity.

The wide distribution of CAD scores for asymptomatic TB is consistent with a wide range of radiological severity, including mild disease (17), and supports the hypothesis that asymptomatic TB is an early phenotype on the TB spectrum (18, 19). qXR CAD sensitivity in both Cohorts appeared marginally higher than human readers, with a corresponding minor decrease in specificity, consistent with other studies in which CAD outperformed human readers (20, 21). However, when the qXR threshold was adjusted to match sensitivity for asymptomatic TB with that observed for human readers in the Screening Cohort, the apparent difference in specificity between qXR CAD and human readers disappeared, suggesting qXR CAD performs similarly to human readers for asymptomatic TB overall. By contrast, in the Triage Cohort, the corresponding threshold-adjusted qXR CAD specificity for symptomatic TB was significantly superior to human readers, which might be expected since qXR CAD was trained on symptomatic TB.

Several CAD platforms have been reported to have moderate to high diagnostic accuracy for TB (21, 22). One study reported an AUC of 0.82 for qXR and DELFT, with lower AUC in a range of 0.50 to 0.73 for other platforms (23). Another study reported AUC of 0.90 for Lunit and Nexus; 0.80 to 0.90 for qXR, InferRead, Xvision, JF CXR-2 and ChestEye; while XrayAME, RADIFY and TiSepX-TB showed AUC less than 0.80 (24). These platforms did not evaluate CAD performance for asymptomatic TB. Further, effectiveness of CAD screening does not depend on performance across the full range of threshold scores reflected by the AUC. Rather, it depends on performance in a narrow band of threshold scores selected to achieve a target sensitivity, with acceptable specificity, for a particular use-case. This point is illustrated by the observation that the AUC for qXR CAD was similar for asymptomatic TB in the Screening Cohort and symptomatic TB in the Triage Cohort, but application of the same 0.5 threshold score resulted in significantly lower sensitivity for asymptomatic TB (69.2%), compared to symptomatic TB (83.6%). A case-control analysis of 1,439 participants selected from five active case-finding studies, some of which used prevalence survey methodology or included high TB risk groups, reported CAD4TB (v7) sensitivity of 61.4% for asymptomatic TB at the manufacturer’s recommended threshold score (50), which is consistent with our findings (25).

We have shown that neither human readers, nor qXR CAD using the manufacturer’s 0.5 threshold score, met the minimal WHO TTP for a high-sensitivity (90%), high-specificity (80%) screening test for asymptomatic TB. It is possible that asymptomatic TB detected through HHC screening is an earlier/milder phenotype than asymptomatic TB in other community settings, but our findings are consistent with the analysis of five active case-finding studies, which showed that dCXR screening with CAD4TB (v7) performed poorly in people who were older, living with HIV, had prior TB, or were asymptomatic; and failed to meet the WHO TPP for a TB screening test (25). Our findings are also consistent with a study of qXR CAD in household contacts, which reported 71% sensitivity and 91% specificity for prevalent TB, although not disaggregated for asymptomatic disease (26). A study of active TB case-finding in the Philippines reported 97% sensitivity and 13% specificity for asymptomatic TB, but these estimates may be subject to selection bias for more severe disease arising from prevalence survey methodology (13).

Our findings raise the question of whether TPP criteria are perhaps too stringent for community screening of asymptomatic TB. Kendall *et al* highlighted the limitations of evaluating TB diagnostics based solely on sensitivity and specificity; and argue that this approach prioritises population-level accuracy without accounting for heterogeneity in the TB disease spectrum (27). Use of varying CAD thresholds has been suggested for different populations (28), recognising that the sensitivity and specificity needed for effective asymptomatic TB screening may differ from that needed to triage symptomatic TB (29). However, the use of varying CAD thresholds for different use-cases would complicate guidelines, training and implementation in the field; and would make direct comparison of CAD performance virtually impossible across different settings and risk groups. We have shown that the qXR CAD threshold required to achieve 90% sensitivity for asymptomatic TB in the Screening Cohort was very low (0.007), compared to that required for symptomatic TB in the Triage Cohort (0.13). It is also apparent that if the qXR CAD threshold were artificially lowered to achieve 90% sensitivity for asymptomatic TB among household contacts in this study, the resultant very low specificity (18.9%) would no longer be feasible for a community-based TB screening programme.

The large proportion of individuals with prior TB among household contacts (20%) and clinic attendees (35%) might have influenced our findings. The overall discriminatory performance of qXR CAD for asymptomatic and symptomatic TB in the Screening Cohort was similar regardless of prior TB history. However, discriminatory performance of qXR CAD for symptomatic TB in the Triage Cohort was significantly better among those without prior TB, compared to those with prior TB, suggesting that residual post-TB radiological changes may impact discriminatory performance, similar to reports from other studies (30, 31). The radiological features and extent of disease associated with asymptomatic TB are the subject of ongoing analysis of data from the RePORT South Africa network.

We suggest that programmatic expectations for community-based dCXR CAD screening should be modelled on comparable data from community-based studies of asymptomatic TB. Our findings among household contacts in South African communities require validation in other settings, but the data raise questions about the impact of mass dCXR CAD screening to detect asymptomatic TB. We stress that these data should not discourage investment in community-based dCXR CAD screening programmes, which would be expected to detect more than two-thirds of people with undiagnosed asymptomatic TB. However, such a programme would not realise its full potential to identify all ‘missing’ TB cases, since dCXR CAD screening with the modest sensitivity observed in this study would leave almost one-third of asymptomatic TB cases undetected, and untreated, with the potential to continue community Mtb transmission and hinder TB control efforts (32).

The strengths of our study include the large number of participants enrolled into two parallel cohorts using a single protocol, at several diverse geographical sites in South Africa, making the findings more generalizable to other high TB burden countries. To our knowledge this is the first study to directly compare the yield of community-based screening for asymptomatic TB vs facility-based triage of symptomatic TB, using parallel dCXR evaluation by human readers blind to TB status and a CAD platform, coupled with standardised TB investigation, universal sputum testing and microbiologically-confirmed TB endpoints.

There were also limitations, including that some symptomatic participants in the Triage Cohort (16.5%) did not have a dCXR available for review. However, the large number of symptomatic TB cases in the Triage Cohort was more than sufficient for comparison with the Screening Cohort, in which asymptomatic TB cases occurred less frequently. A small percentage (8%) of original CXR from symptomatic participants in the Triage Cohort were obtained in analogue format and subsequently digitalised, potentially reducing image quality and discriminatory performance, but dCXR from asymptomatic participants in the Screening Cohort were not affected. Human dCXR readers were site investigators with the clinical training and expertise expected of research clinicians in a TB-endemic setting. However, this level of expertise might be considered advanced for some high-burden countries in which dCXR TB screening might be implemented. Our study used a single CAD platform, qXR v3.0, and it is possible that other CAD platforms might have performed better or worse for community-based screening of asymptomatic TB (11, 23, 24). However, qXR CAD software was selected because it was reported to have among the best discriminatory performance of the six WHO-endorsed CAD platforms (11); and of all 12 CAD platforms evaluated by two recent studies (23, 24).

## Conclusion

Sensitivity of community-based dCXR screening for asymptomatic TB among household contacts was low, compared to historical benchmarks for symptomatic TB, but approached 70% with CAD. qXR CAD sensitivity appeared marginally higher than human readers, with a corresponding minor reduction in specificity, but neither dCXR screening approach met the WHO TPP targets for a high sensitivity (90%), high specificity (80%) TB screening test. Although implementation of national dCXR CAD community screening programmes would be expected to detect more than two-thirds of individuals with undiagnosed asymptomatic TB, the significant proportion of cases that would remain undetected, and untreated, might be sufficient to allow ongoing community Mtb transmission and hinder elimination efforts.

## Contributors

TJS, TRS, and MH conceived the study and sourced funding. GW, TJS, TRS, and MH wrote the parent protocol and provided study oversight. SN wrote the analytic protocol with supervision by MH. KH provided operational and project management. SN, MT, TM, SCM, FM, JS, NT, STM, FN AKKL, KD, TP, NM, AL, and BF were responsible for site-level activities, including recruitment, clinical management, and data collection. HM, YFvdH, FM, RP, KH, and KS cleaned and verified underlying data. SN analysed and interpreted the data and wrote the first draft of the manuscript under supervision from HM and MH. All authors reviewed and approved the presentation and interpretation of the final manuscript for submission. RePORT South Africa study team members are listed in the Supplement.

## Conflict of interest declaration

SCM reports research grant support from the Gates Foundation, the South African Medical Research Council (SAMRC), and CRDF Global to the University of Cape Town, and honoraria from CRDF Global for reviewing grant applications. NM reports grant support from SAMRC and CRDF Global. TRS reports grant support from CRDF Global. MH reports institutional research grants to University of Cape Town. All other authors declare no competing interests.

## Data availability statement

The data may be made available upon request from the principal investigator and corresponding author Prof Mark Hatherill, subject to approval of a concept proposal by the RePORT South Africa network.

## Acknowledgements and funding sources

We are grateful to the study participants and their communities in which this research was conducted. This research was supported by the RePORT South Africa network with funds received from CRDF Global (University of Cape Town, G-DAA3-19-66875-1; University of Cape Town Lung Institute, G-DAA9-20-66917-1; Vanderbilt University Medical Center, G-DAA9-20-66870-1; Stellenbosch University, G-DAA9-20-66918-1; Africa Health Research Institute G-202403-71799; University of Pretoria, G-DAA9-20-66880-1 and Wits Health Consortium, G-DAA9-20-66878-1); the US National Institutes of Health (NIH; Stellenbosch University, U01AI152075), and the South African Medical Research Council (SAMRC). The content and findings reported are the sole deduction, view and responsibility of the researcher and do not reflect the official position and sentiments of the SAMRC or the NIH.

## Notes

### Author Declarations

This analysis was approved by the Human Research Ethics Committee of the University of Cape Town. The RePORT South Africa study protocol was approved by the institutional ethics committees of all participating sites.

