## Supplement for "Performance of Human and Computer-aided Evaluation of Digital Chest Radiography for Community-based Screening of Asymptomatic Tuberculosis"

**Author affiliations:**

### RePORT South Africa team members

| **Name** | **Affiliation** |
| --- | --- |
| Pattamukkil Abraham | Perinatal HIV Research Unit, University of Witwatersrand |
| Denis Awany | South African Tuberculosis Vaccine Initiative, University of Cape Town |
| Cynthia Baard | University of Cape Town Lung Institute |
| Zainab Baig | Africa Health Research Institute |
| John Belisle | Colorado State University |
| Rebecca Berhanu | Vanderbilt Tuberculosis Center, Vanderbilt University Medical Center |
| Nicole Bilek | South African Tuberculosis Vaccine Initiative, University of Cape Town |
| Gerard Cangelosi | University of Washington |
| ACE Carstens | Department of Biomedical Sciences, Stellenbosch University |
| Kevyna Chetty | Africa Health Research Institute |
| Yolundi Cloete | South African Tuberculosis Vaccine Initiative, University of Cape Town |
| Rodney Dawson | University of Cape Town Lung Institute |
| Marwou de Kock | South African Tuberculosis Vaccine Initiative, University of Cape Town |
| Gareta Dickman | Africa Health Research Institute |
| Charity Dire | Perinatal HIV Research Unit, University of Witwatersrand |
| Karen Dobos | Colorado State University |
| Stephany Norah Duda | Vanderbilt Tuberculosis Center, Vanderbilt University Medical Center |
| Mzwandile Erasmus | South African Tuberculosis Vaccine Initiative, University of Cape Town |
| Aliasgar Esmail | Division of Pulmonology, Department of Medicine, Groote Schuur Hospital and University of Cape Town Lung Institute |
| Marika Flinn | Department of Biomedical Sciences, Stellenbosch University |
| Travis Harris | Vanderbilt Tuberculosis Center, Vanderbilt University Medical Center |
| Andriette Hiemstra  Bernadine Fransman | Department of Biomedical Sciences, Stellenbosch University  Department of Biomedical Sciences, Stellenbosch University |
| Shameem Jaumdally | University of Cape Town Lung Institute |
| Ryan Johnson | University of Cape Town |
| Farina Karim | Africa Health Research Institute |
| Masooda Kaskar | South African Tuberculosis Vaccine Initiative, University of Cape Town |
| Nobulumko Khomba | South African Tuberculosis Vaccine Initiative, University of Cape Town |
| Thandeka Khoza | Africa Health Research Institute |
| Léanie Kleynhans | Department of Biomedical Sciences, Stellenbosch University;  Mater Research Institute – The University of Queensland |
| Tahira Kootbodien | University of Cape Town |
| Andrea Kotze | University of Cape Town Lung Institute |
| Belinda A Kriel | Department of Biomedical Sciences, Stellenbosch University |
| Ané Kruger | Department of Biomedical Sciences, Stellenbosch University |
| Alasdair Leslie | Africa Health Research Institute |
| Lorraine Lichakane | Perinatal HIV Research Unit, University of Witwatersrand |
| Ilze Louw | Department of Biomedical Sciences, Stellenbosch University |
| Simbarashe Mabwe | South African Tuberculosis Vaccine Initiative, University of Cape Town |
| Candice MacDonald | Department of Biomedical Sciences, Stellenbosch University |
| Lindiwe Madziwa | Africa Health Research Institute |
| Lebohang Makhethe | South African Tuberculosis Vaccine Initiative, University of Cape Town |
| Sandisiwe Mangali | South African Tuberculosis Vaccine Initiative, University of Cape Town |
| Neil Martinson | Perinatal HIV Research Unit, University of Witwatersrand |
| Linda Mbuthini | University of Cape Town |
| Carolina Mehaffy | Colorado State University |
| Thabang Moloja | Perinatal HIV Research Unit, University of Witwatersrand |
| Angelique Mouton | South African Tuberculosis Vaccine Initiative, University of Cape Town |
| Mbusiseni Ngema | Perinatal HIV Research Unit, University of Witwatersrand |
| Hlengiwe Nkambule | South African Tuberculosis Vaccine Initiative, University of Cape Town |
| Onke Nombida | South African Tuberculosis Vaccine Initiative, University of Cape Town |
| Fajwa Opperman | South African Tuberculosis Vaccine Initiative, University of Cape Town |
| Gregory Ording-Jespersen | Africa Health Research Institute |
| Kennedy Otwombe | Perinatal HIV Research Unit, University of Witwatersrand |
| Tracy Richardson | Department of Biomedical Sciences, Stellenbosch University |
| Carmen Segelaar | South African Tuberculosis Vaccine Initiative, University of Cape Town |
| Jane Shaw | Department of Biomedical Sciences, Stellenbosch University |
| Kimberly Shelton | Colorado State University |
| Theresa Smit | Africa Health Research Institute |
| Bronwyn Smith | Department of Biomedical Sciences, Stellenbosch University |
| Candice Snyders | Department of Biomedical Sciences, Stellenbosch University |
| Marcia Steyn | South African Tuberculosis Vaccine Initiative, University of Cape Town |
| Sara Suliman | University of California San Francisco |
| Floris Swanepoel | Perinatal HIV Research Unit, University of Witwatersrand |
| Lorraine Thobakgale | Perinatal HIV Research Unit, University of Witwatersrand |
| Susanne Tonsing | Department of Biomedical Sciences, Stellenbosch University |
| Petrus Tyambetyu | South African Tuberculosis Vaccine Initiative, University of Cape Town |
| Habibullah Valley | South African Tuberculosis Vaccine Initiative, University of Cape Town |
| Lyle van de Berg | Perinatal HIV Research Unit, University of Witwatersrand |
| Gian van der Spuy | Department of Biomedical Sciences, Stellenbosch University |
| Ilana R van Rensburg | Department of Biomedical Sciences, Stellenbosch University |
| Johanna E van Rooyen | South African Tuberculosis Vaccine Initiative, University of Cape Town |
| Ashley Veldsman | South African Tuberculosis Vaccine Initiative, University of Cape Town |
| Lindsay Wilson | University of Cape Town |
| Rachel C Wood | University of Washington |
| Lesley Workman | University of Cape Town |
| Heather Zar | Department of Paediatrics and Child Health, University of Cape Town |

### Table S1: Research in context summary findings.

| **Study Year** | **Setting** | **Population** | **CAD software** | **Screening approach** | **CAD performance for**  **Asymptomatic TB** | | | **Comment** | **Met WHO TPP (sensitivity 90%, specificity 80%)** |
| --- | --- | --- | --- | --- | --- | --- | --- | --- | --- |
|  |  |  |  |  | **AUC**  **(95% CI)** | **Sensitivity (95% CI)** | **Specificity**  **(95% CI)** |  |  |
| Mungai et al, (2022) (1) | 2016 Kenya National Tuberculosis Prevalence Survey | All persons 15 years and older, N=61,848.  Asymptomatic TB cases not reported separately. | CAD4TB  v6 (threshold 55) | Community screening.  Used prevalence survey methodology. Only symptom-positive and/or CXR-positive individuals underwent sputum testing. Symptom-negative, CXR-negative individuals not tested. | Not reported | Not reported | Not reported | Did not report AUC, sensitivity or specificity for asymptomatic TB separately. | NA |
| Marquez et al, (2025) (2) | Four regions of the Philippines, May 2021–March 2024 | All persons 15 years and older, N=5,740.  Asymptomatic TB cases n=478 (8.3%) | qXR  (threshold 0.5) | Active case-finding in communities and workplaces; intensified case-finding in hospitals  Used prevalence survey methodology. Only symptom-positive and/or CXR-positive individuals underwent sputum molecular WHO-recommended rapid diagnostic (mWRD). Symptom-negative, CXR-negative individuals not tested. | AUC  0.79  (0.77–0.81) | Sensitivity 97.1% (96.5–97.6) | Specificity 13.2% (12.2–14.3) | Potential selection bias toward more severe disease in hospitalised patients; and due to exclusion of symptom-negative, CXR-negative participants from diagnostic sputum testing. | No |
| Macpherson et al, (2025) (3) | South Africa, November 2014–May 2021 | Household contacts 18 years and older, N=483.  Asymptomatic TB cases in the biomarker subgroup (N=247)  n=6 (2.4%) | qXR  v3.0  (threshold 0.5)  CAD4TB  v7.0  (threshold 50)  Lunit INSIGHT  v3.4.111  (threshold 0.15) | Household contact tracing of rifampicin-resistant TB patients.  Used universal TB screening. Sputum tested from all participants, regardless of symptom or CXR status. | qXR  v3.0  AUC 0.90  (0.74–1)  CAD4TB  v7  AUC 0.95 (0.89–1)  Lunit  v3  AUC 0.98 (0.93–1) | Not reported | Not reported | Did not report sensitivity or specificity for asymptomatic TB separately. | NA |
| Fehr et al, (2021) (4) | South Africa, May 2018–May 2019 | All persons 15 years and older, N=9,914.  Asymptomatic TB cases  n=79 (0.8%) | CAD4TB  v5 and v6 (threshold 25) | Community screening  (mobile health camps)  Used prevalence survey methodology. Only symptom positive and CXR-positive individuals underwent sputum testing based on CAD4TB v5 score threshold of 60. Symptom-negative, CXR-negative individuals were not tested. | Not reported | Not reported | Not reported | Did not report AUC, sensitivity or specificity for asymptomatic TB separately. | NA. |
| Scott et al, (2025) (5) | South Africa, November 2016–August 2023 | Case-control study of pooled datasets from five active case-finding studies in South Africa.  N=1,439 TB cases and controls 15 years and older, selected from a total study population of 20,770 (6.9%).  Asymptomatic TB cases  n=285 (1.4%) | CAD4TB v7 (threshold  50) | Community-based, active case-finding, including high-risk groups.  Two studies used prevalence survey methodology. Only symptom-positive and/or CXR-positive individuals underwent sputum testing. Symptom-negative, CXR-negative individuals were not tested.  Three studies performed diagnostic sputum testing in symptom-positive participants and in those with an additional risk factor for TB. Symptom-negative individuals without a TB risk factor were not tested. Two of these studies performed CXR only in individuals with a TB risk factor. | AUC  0·79 (0·76–0·82) | Sensitivity  61.4% (57.9-64.9) | Specificity  86.7% (83.2-90.2) | Case-control analysis included <5% of all participants without TB.  Potential selection bias toward more severe disease due to exclusion of symptom-negative, CXR-negative participants from diagnostic sputum testing (2 studies); inclusion of high-risk groups, such as PLWH and those with previous TB (3 studies); and exclusion of participants without a risk factor from diagnostic sputum testing (2 studies). | No |

CXR, chest radiography. CAD, computer aided diagnostic. AUC, area under the curve. TPP, target product profile. ACF, active case finding.

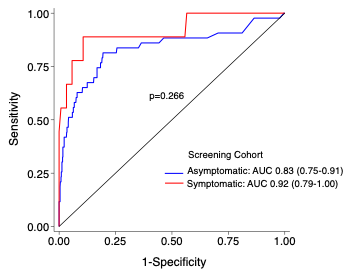

### Figure S1: Discriminatory performance of qXR stratified by symptom status in the Household Contact Screening Cohort.

Receiver operating characteristic (ROC) curves for qXR in symptomatic and asymptomatic participants within the Household Contact Screening Cohort. Area under the curve (AUC) with 95% confidence interval are shown separately for symptom-defined subgroups (n=39 asymptomatic TB cases; and n=716 asymptomatic controls without TB; n=9 symptomatic TB cases and n=122 symptomatic controls without TB). The p-value shown is derived from the DeLong test comparing AUCs between symptomatic and asymptomatic groups.

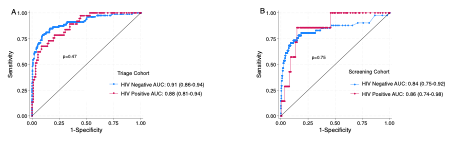

### Figure S2: Discriminatory performance of qXR stratified by cohort and HIV status.

Receiver operating characteristic (ROC) curves for qXR differentiating participants with TB from those without TB in the (**A**) Facility-based Triage and (**B**) Household Contact Screening cohorts, stratified by HIV status. Area under the curve (AUC) with 95% confidence intervals are shown separately for HIV-defined subgroups within each cohort (Triage Cohort: n=129 people living with HIV and n=338 HIV negative; Screening Cohort: n=149 people living with HIV and n=737 HIV negative). The p-values shown are derived from the DeLong test comparing AUCs between people living with HIV and HIV-negative groups within each cohort.
